# COVID-19 Vaccination acceptance in the canton of Geneva: A Cross-Sectional Population-Based Study

**DOI:** 10.1101/2021.07.05.21260024

**Authors:** Ania Wisniak, Hélène Baysson, Nick Pullen, Mayssam Nehme, Francesco Pennacchio, María-Eugenia Zaballa, Idris Guessous, Silvia Stringhini

## Abstract

**Objective:** This study aimed to assess acceptance of COVID-19 vaccination as well as its sociodemographic and clinical determinants in a general population sample three months after the launch of the vaccination program in Switzerland.

**Methods:** In March 2021, an online questionnaire on vaccination acceptance was proposed to adults included in a longitudinal cohort study of previous population-based serosurveys carried out in the canton of Geneva. Questions were asked about COVID-19 vaccination acceptance, reasons of acceptance or refusal, and attitudes about vaccination in general. Data on demographic (age, sex, education, income, professional status, living conditions) and health-related characteristics (having a chronic disease, COVID-19 diagnosis, smoking status) were assessed at inclusion in the cohort (December 2020).

**Results:** Overall, 4’067 participants (completion rate of 77.4%) responded to the survey between March 17 and April 1, 2021. The mean age of respondents was 53.3 years and 56.0% were women. Most had completed tertiary education (64.7%) and over 60% were currently professionally active. At the time of the survey, 17.2% of respondents had already been vaccinated with at least one dose or had made an appointment to get vaccinated, and an additional 58.5% intended or rather intended to get vaccinated. The overall acceptance of COVID-19 vaccination was 75.7%, with a higher acceptance among men compared to women, older adults compared to younger adults, high-income individuals compared to those with a low income, participants living in urban and semi-urban areas compared to rural, and retirees and students compared to employed individuals. Acceptance was lower among individuals having completed apprenticeships and secondary education compared to those with tertiary education. The most common reasons reported by participants intending to get vaccinated were the desire to ‘get back to normal’, to protect themselves, their community and/or society, and their relatives or friends against the risk of infection by SARS-CoV-2, as well as the desire to travel. Less than half (45.6%) of participants having children were willing or rather willing to have their children vaccinated against COVID-19 if it were recommended by public health authorities.

**Conclusion:** Although our study found a 75.7% acceptance of COVID-19 vaccination, there were noticeable socio-demographic disparities in vaccination acceptance. These data will be useful for public health measures targeting hesitant populations when developing health communication strategies. These results will be updated over time with a new release of the survey in autumn 2021.

## Introduction

In response to the COVID-19 pandemic, unprecedented global efforts have enabled the development of several safe and effective vaccines only one year after the first COVID-19 case was diagnosed in Wuhan, China. Worldwide, the first COVID-19 vaccines tested in phase 3 trials were commercialized at the beginning of December 2020, with the messenger RNA-based Comirnaty® vaccine of Pfizer/BioNTech^1^ being the first authorized in the United Kingdom on December 3, 2020. In addition to manufacturing and logistical challenges, vaccination campaigns worldwide have been challenged by diffuse distrust of the population regarding the safety and efficacy of these novel vaccines.2–6

Vaccine hesitancy fueled by misinformation campaigns has often been a threat to sufficient vaccine coverage over past decades, sometimes leading to resurgence of vaccine-preventable diseases.7–9 This has led the World Health Organization to recognize vaccine hesitancy as a major threat to global health in 2019.10 International efforts to urgently deliver a safe and effective vaccine against COVID-19 have been faced with a growing anti-vaccination movement amplified by social media since the early phases of the pandemic, with the potential to negatively impact vaccination uptake in populations exposed to these campaigns.2,5,11–13

By December 23, 2020, the date of the launch of COVID-19 vaccination, Switzerland had reported 4’896 confirmed cases/100’000 inhabitants and 6’406 deaths since the beginning of the pandemic.^14^ In addition to the direct health consequences of COVID-19, social distancing measures and closure of non-essential services have led to negative social, psychological and economic consequences. In the canton of Geneva, a population-based serological survey has shown that by the end of December 2020, 21.1% of the canton’s population had been infected with SARS-CoV-2 since the start of the pandemic^15^, suggesting a relatively slow rise of population immunity if social distancing measures preventing the collapse of health care systems were to be pursued. At the time of our survey, during the last two weeks of March 2021, two messenger RNA-based vaccines were available in Switzerland – the Comirnaty® (BNT162b2) vaccine of Pfizer/BioNTech^1^ and the COVID-19 vaccine (mRNA-1273) of Moderna^16^ vaccines. At that time in the canton of Geneva, vaccination priority was given to individuals aged 65 years and older, individuals deemed ‘particularly vulnerable to COVID-19’, as well as health workers in close contact with at-risk patients.^17^

Reaching sufficient coverage, however, is in large part dependent on the population’s willingness to get vaccinated. A national survey conducted in Switzerland shortly before the arrival of the first vaccines on the market revealed that only 56% of respondents were likely to accept vaccination against COVID-19, with a lack of trust in the security of the vaccines being the main reason for refusal.18 Furthermore, previous studies have shown that vaccine hesitancy was associated to socio-demographic factors such as younger age, female gender, lower income and lower education.^2,19,20^ In order to address vaccine hesitancy in a comprehensive way and deliver targeted interventions, reasons for accepting or refusing vaccination, as well as associated socio-economic factors should be explored in a regional context, as results found in other countries cannot be extended to all populations, due to cultural, political and organizational factors influencing vaccination acceptance. In addition, taking into account people’s positive or negative emotions about vaccination is essential to developing effective communication campaigns.^21^

The aim of our study was 1) to assess the population’s willingness to get vaccinated against COVID-19 three months after the launch of the vaccination program in Geneva, Switzerland, 2) to explore individuals’ attitudes towards COVID-19 vaccination and their reasons for accepting or declining the vaccination, and 3) to describe associations between socio-economic or health-related factors and vaccine hesitancy.

## Methods

### Study design, setting and sample

This population-based cross-sectional study was embedded in a longitudinal digital cohort study called Specchio-COVID19, which was launched in December 2020 to follow up over time participants of serosurveys conducted in the canton of Geneva.^22^ Serosurvey participants were randomly selected from the general population at two time points: 1) between April and June 2020, participants were enrolled from a previous general health survey representative of the population of the Canton of Geneva aged between 20 and 75 years old^23^, and 2) between November and December 2020, participants were randomly selected from registries of the Canton of Geneva stratified by age and sex^15^.

After a baseline serologic test, participants were invited to join the Specchio-COVID19 study, which consists in a long-term follow-up by collecting data through regular on-line questionnaires and serological follow-up. Upon registration, an initial questionnaire assessed socio-demographic and lifestyle characteristics and general health-related data. Self-reported SARS-CoV-2 infections and risk perception of COVID-19 are updated through monthly questionnaires. The questionnaire designed for this study was sent out to participants on March 17, 2021, with a reminder sent two weeks later.

### Data collected in the COVID-19 vaccination questionnaire

The “vaccination” questionnaire was based on a literature review and was validated by public health experts and physicians. Part of the content was developed in the framework of the Corona Immunitas research program, a national program aiming to coordinate regional SARS-CoV-2 seroprevalence studies across Switzerland.^24^ The questionnaire measured COVID-19 vaccination status, intention to get vaccinated, reasons to get vaccinated, reasons for refusing vaccination, vaccination-related beliefs (e.g., perceived efficacy, perceived safety, preference for natural immunity), perceived utility of COVID-19 vaccination, willingness to vaccinate one’s children against COVID-19, and attitude towards vaccination in general. The questionnaire also included three questions from a French study on vaccination hesitancy^25^ adapted from the World Health Organization’s Strategic Advisory Group of Experts (SAGE) definition of vaccine hesitancy^26^, general attitudes regarding vaccination, and trust in public health authorities, pharmaceutical companies, scientists and researchers. These questions are described in more detail in the supplementary materials. Two questions were additionally asked on the perception of immunity certificates for COVID-19, for which analyses were conducted and described in a separate paper submitted to the same journal.

Vaccination intention was defined as the combined answer to the following two questions: “Were you already vaccinated against COVID-19? (Yes, No, Scheduled appointment)” and “Do you intend to get vaccinated once you will be eligible for vaccination against COVID-19? (Yes, rather yes, rather no, no, does not know)”. Answers “Yes”, “Scheduled appointment” to the first question and answers “Yes” and “Rather yes” to the second question were later combined as willingness to get vaccinated. Answers “No” and “Rather no” to the second question were defined as no willingness to get vaccinated.

We constructed the variable ‘Vaccine hesitancy’ (hesitant/not hesitant) based on the SAGE definition, categorizing as ‘hesitant’ participants who had at some point refused vaccination and/or delayed vaccination and/or accepted vaccination despite doubts on its effectiveness. Those who answered ‘no’ to all three questions were considered ‘not vaccine hesitant’.

Previous SARS-CoV-2 infection was defined as being either SARS-CoV-2 seropositive or having self-reported a positive PCR or antigenic test for SARS-CoV-2 in one of the monthly surveys.

Education was categorized as follows: 1) compulsory education or no formal education, 2) apprenticeships, 3) secondary school and specialized schools, and 4) tertiary education including universities, higher professional education, and doctorates.

Income was categorized as ‘low’ (below the first quartile of the general population of the canton of Geneva), ‘medium’ (between the first and third quartiles) or ‘high’ (above the third quartile) taking into account self-reported household income from the baseline questionnaire, as well as household composition (living alone with or without children, in a relationship with or without children, or in a shared apartment with other adults), and according to household income statistics for the same household composition categories within the canton of Geneva. ^27^

### Statistical analysis

Descriptive analyses included percentages with comparisons using chi-square tests for categorical variables. P-values were considered significant at p<0.05.

Logistic regression analyses were conducted to assess the associations of demographic and health-related factors with COVID-19 vaccination intention. Simple univariate logistic models were run for all of the following variables individually: sex, age, education, household income, residential area, employment status, living conditions, having a chronic disease, smoking status, previous SARS-CoV-2 infection, perception of COVID-19 severity and contagiousness, and vaccine hesitancy. For each variable we also ran multivariable logistic regressions adjusting for age, sex, education and income. The intersex category as well as the “non available” (NA) categories for all variables were excluded from the logistical regression analysis due to low counts. Odds ratios and confidence intervals were calculated through exponentiation of estimated coefficients. Statistical significance was taken at the level of p<0.05 *a priori*. All analyses were conducted using R 4.0.3 (R Foundation for Statistical Computing, Vienna, Austria).

### Ethical Considerations

All participants of the Specchio-COVID19 digital platform provided informed and written consent upon enrolment in the study. Ethical approval for the study was obtained from the Cantonal Research Ethics Commission of Geneva, Switzerland (project number 2020-00881).

## Results

### Descriptive characteristics of the sample

From the original 8’904 adult serosurvey participants invited to be followed-up longitudinally, 5’282 enrolled in the digital cohort (participation rate 59.3%, not taking into account unreachable participants due to false email addresses), among which 30 withdrew their participation prior to the vaccination survey. Overall, 4’067 participants (completion rate of 77.4%) responded to the vaccination survey between March 17 and April 1, 2021 (the study flow chart is presented in supplementary materials, figure S1). The mean age of participants was 53.3 years (+/-14.4 years standard deviation) and 56.0% were women. Most had completed a tertiary education (n=2’631; 64.7%) and over 60% were currently professionally active (as an independent or an employee). Overall characteristics are presented in supplementary materials (Table S1). In comparison with the general population in the Canton of Geneva, our sample had older participants (27.9% individuals aged 18-34 years in Geneva vs. 10.4% in our sample) and a higher education level (64.9% higher education in our sample vs. 39.9% in the Geneva population) (Supplementary materials, Table S2).

Compared to non-respondents, participants responding to the vaccination survey were older (mean age 53.3 +/- 14.4 years vs. 43.8 +/- 14.4 years), more highly educated (64.7% vs. 63.2% had tertiary education, 3.9% vs. 6.8% had no formal education, p<0.001), had a higher income (12.9% vs. 16.3% had low income, 64.8% vs. 55.9% middle or high income, p<0.001), were more frequently retired (25.9% vs. 9.8%) and less frequently students (4% vs. 12%, p<0.001) and were less frequently current smokers (14.9% vs. 19.2%, p<0.001). Socio-demographic and health-related characteristics of survey respondents compared to non-respondents are presented in supplementary materials (Table S1).

### Vaccination status and intention

At the time of the survey, 17.2% of respondents had already been vaccinated with at least one dose or had made an appointment to get vaccinated. Moreover, 58.5% of participants intended or rather intended to get vaccinated, while only 13.8% did not or rather did not intend to get vaccinated, and 10.4% did not know if they intended to get vaccinated (Figure 1).

**Figure 1.**
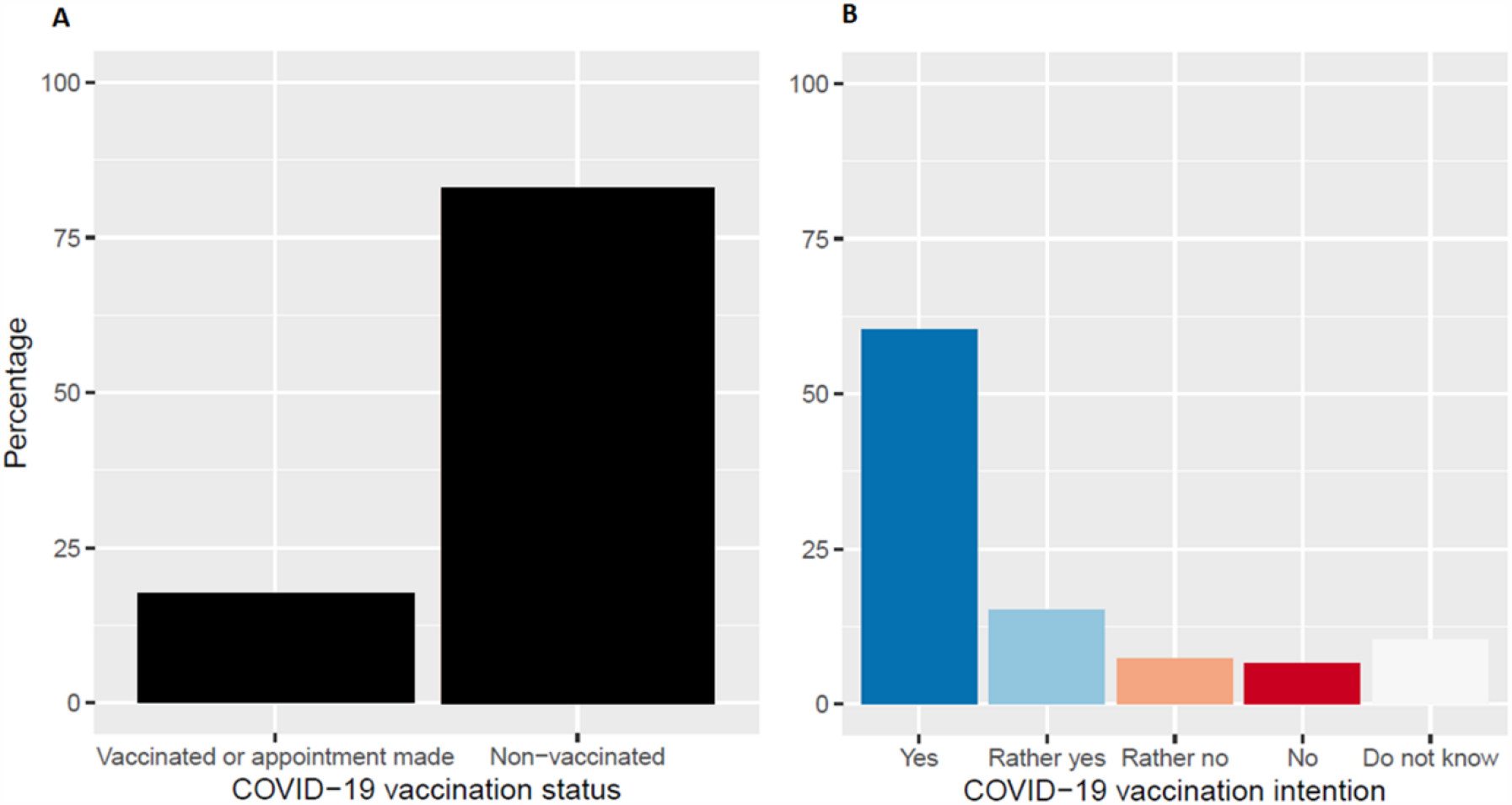
**A**. Proportion of participants vaccinated/with appointment for vaccination versus participants not vaccinated against COVID-19. **B**. Intention to get vaccinated against COVID-19; «Yes» combines those willing to get vaccinated and those already vaccinated or who have an appointment.

### General perception of COVID-19 vaccination usefulness

A large majority (92.3%) agreed or rather agreed that COVID-19 vaccination was an important step to end the pandemic. When stratified by vaccination intention, those willing to or already vaccinated agreed the most with this statement (99.4%), although even those not intending to get vaccinated acknowledged the importance of vaccination at a majority (57.1%) (Figure 2A). Similarly, a majority of participants (78.5%) considered that vaccinated individuals should continue following preventive measures such as wearing face masks. Individuals willing to or already vaccinated were more likely to agree with this statement (81.9%) when compared to those not willing to get vaccinated (64.6%) (Figure 2B).

**Figure 2.**
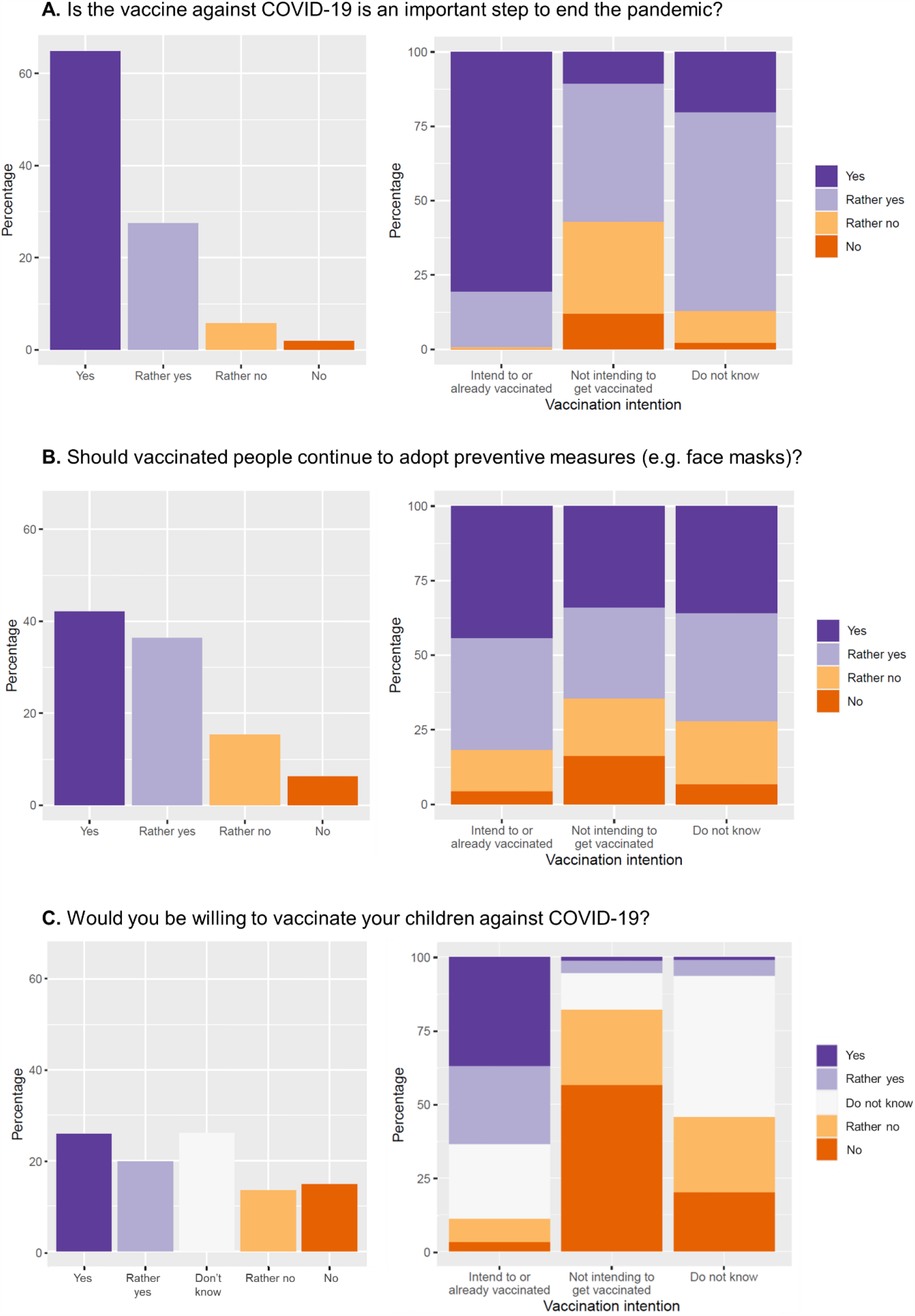
**A**. Proportion of participants agreeing that the COVID-19 vaccine is an important step to end the pandemic, in the overall sample (left) and stratified by vaccination intention (right). **B**. Proportion of participants agreeing that vaccinated individuals should continue to adopt preventive measures, in the overall sample (left) and stratified by vaccination intention (right). **C**. Proportion of participants willing to vaccinate their children, in the sample of participants with children under 18 years old (left) and stratified by vaccination intention (right).

### Willingness to vaccinate children

Participants with children under the age of 18 (N=1’339) were asked whether they would be willing to have their children vaccinated against COVID-19 if it was recommended by public health authorities. Less than half (45.6%) agreed or rather agreed, and approximately one quarter did not know (Figure 2C). When stratified by vaccination intention for oneself, those intending or rather intending to get vaccinated were mainly willing to have their children vaccinated (63.6%), while among participants not intending to get vaccinated or not yet sure, they were only 5.6% and 6.5%, respectively. Importantly, a high proportion of parents intending to get vaccinated (25.3%) and not yet sure about their intention (47.8%) were still undecided regarding vaccination of their children against COVID-19. These results were also stratified by parents’ education level and children’s age (the youngest child’s age was considered for parents with more than one child), showing the highest willingness rate for children’s vaccination among the most (50.5%) and the least (46.2%) educated, and an apparent gradient in willingness with increasing children’s age from 6 years old (between 38.6% for children aged 6 to 10, to 55.9% for children aged 16 to 18). These results are detailed in supplementary materials (Table S3).

### Reasons for Covid-19 vaccine acceptance and refusal

Main reasons for intending to get vaccinated and for refusing vaccination are listed in Table 1. The most common reasons reported by participants were the desire to ‘get back to normal’ (78.4%), and to protect themselves (75.4%), as well as their community and/or society (70.1%) against the risk of infection by SARS-CoV-2.

**Table 1.** **1a)** Reasons for accepting or refusing vaccination, and **1b)** reasons that might change participants’ minds.

| **1a. Reasons to get vaccinated (if vaccination intention=yes/rather yes)**<br>**(N=2'379\*)** | **N (%)** |
| --- | --- |
| Desire to 'get back to normal' | 1866 (78.4) |
| Protect myself against infection | 1794 (75.4) |
| Protect the community/society | 1667 (70.1) |
| Desire to travel | 1637 (68.8) |
| Protect those close to me | 1510 (63.5) |
| Adherence to public health recommendations | 1145 (48.1) |
| Living or working with vulnerable people | 311 (13.1) |
| At risk of infection at the workplace | 280 (11.8) |
| At risk of complications due to age | 206 (8.7) |
| At risk of complications due to health state | 138 (5.8) |
| Vaccine recommended by healthcare professional | 107 (4.5) |
| At risk for other reasons | 72 (3) |
| Employer required vaccination | 40 (1.7) |
| **Reasons for refusing vaccination (if vaccination intention=No/rather no)**<br>**(N=562)** | **N (%)** |
| Prefer waiting | 303 (53.9) |
| Waiting for additional information | 162 (53.5) |
| Give priority to more vulnerable people | 73 (24.1) |
| Waiting for more people to get vaccinated | 47 (15.5) |
| Waiting for my serological test result | 13 (4.3) |
| Not afraid of getting infected | 155 (27.6) |
| Not at risk of complications | 88 (56.8) |
| I protect myself | 25 (16.1) |
| I have antibodies against SARS-CoV-2 | 20 (12.9) |
| COVID-19 is a trivial disease | 13 (8.4) |
| Preference for other preventive measures | 154 (27.4) |
| Preference for natural/traditional treatments | 50 (32.5) |
| Preference for natural immunity | 46 (29.9) |
| Preference for other means of protection | 40 (26) |
| Worried or afraid to get vaccinated | 137 (24.4) |
| Afraid of long-term side-effects | 71 (51.8) |
| Vaccine developed too fast | 49 (35.8) |
| Distrust in biological mechanism of the vaccine | 14 (10.2) |
| Feel protected because previously infected by SARS-CoV-2 | 133 (23.7) |
| Believe that vaccine doesn't prevent transmission | 117 (20.8) |
| Against vaccines in general | 76 (13.5) |
| The sanitary situation is improving | 22 (3.9) |
| Can't get vaccinated for medical reasons | 6 (1.1) |
| **1c. If no/rather no, reasons that may change participants' minds (N=562)** |  |
| More reliable information on vaccine's efficacy | 289 (51.4) |
| Scientific results showing low risk of side effects | 284 (50.5) |
| Mandatory vaccination for certain situations (e.g. travel) | 188 (33.5) |
| Deterioration of the sanitary situation | 69 (12.3) |
| Better communication by authorities | 54 (9.6) |
| Many people in Switzerland getting vaccinated | 24 (4.3) |
| Reassuring information in the media or social media | 21 (3.7) |
| Friends or relatives getting vaccinated | 13 (2.3) |
| Will not change mind | 69 (12.3) |

\*Does not include participants who were already vaccinated or who had an appointment to get vaccinated.

Among those not intending to get vaccinated, the most common reason selected was the ‘preference to wait’ by 53.9% of participants. Other common reasons for refusing vaccination were not being afraid of being infected by SARS-CoV-2 (27.6%), the preference for other preventive measures (27.4%), worry or fear of getting vaccinated (24.4%), feeling protected by a previous infection with SARS-CoV-2 (23.7%) and believing that the vaccine does not prevent transmission of the virus (20.8%). Overall, 13.5% of participants not intending to get vaccinated stated being against vaccines in general.

Participants who did not intend to get vaccinated against COVID-19 were additionally asked which elements would change their minds in favor of vaccination. More than half indicated that more reliable information on vaccine efficacy and scientific results showing low risk of side effects might make them more favorable towards getting vaccinated, and 33.5% reported that making vaccination mandatory in certain contexts (e.g. traveling) would have that effect. Overall, 12.3% of those not willing to get vaccinated stated that they would not change their minds.

### Change in vaccination intention

Overall, in the three months preceding the questionnaire, 21.9% of all participants declared a change in intention to get vaccinated against COVID-19, with most becoming more favorable towards vaccination (19.8%) (Table 2a). Of note, those participants who declared still being ambivalent towards vaccination (‘do not know’) had mostly changed their minds in favor of vaccination (20.5% vs. 5.2%), while those who did not intend to get vaccinated became more or less in favor of vaccination in more equal proportions (5.9% vs. 6.8%, respectively).

**Table 2.**
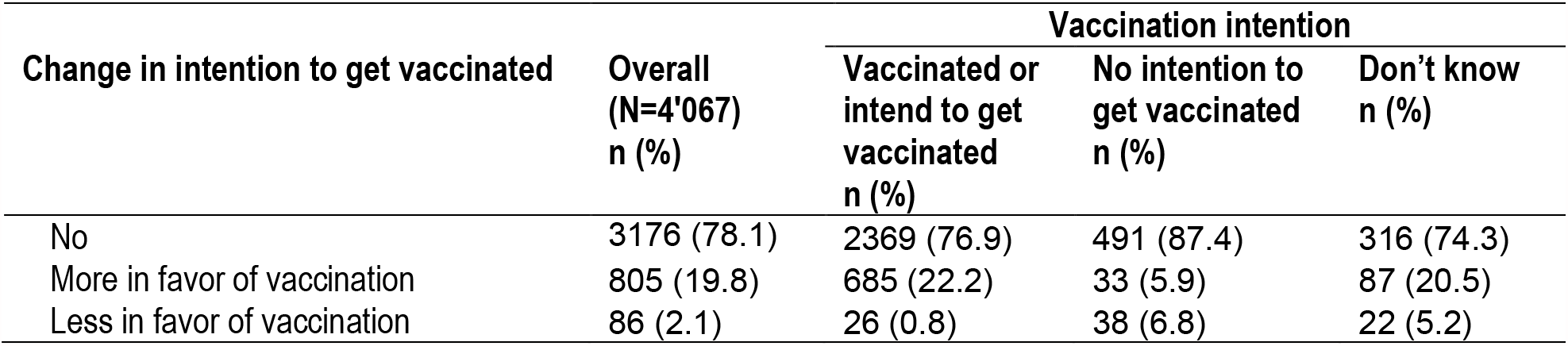
Change in intention to get vaccinated against COVID-19 in the past three months.

| Change in intention to get vaccinated | Overall<br>(N=4'067)<br>n (%) | Vaccination intention |  |  |
| --- | --- | --- | --- | --- |
|  |  | Vaccinated or<br>intend to get<br>vaccinated<br>n (%) | No intention to<br>get vaccinated<br>n (%) | Don't know<br>n (%) |
| No | 3176 (78.1) | 2369 (76.9) | 491 (87.4) | 316 (74.3) |
| More in favor of vaccination | 805 (19.8) | 685 (22.2) | 33 (5.9) | 87 (20.5) |
| Less in favor of vaccination | 86 (2.1) | 26 (0.8) | 38 (6.8) | 22 (5.2) |

Among the participants who changed their mind in the past three months, those more in favor of vaccination indicated the change in the sanitary situation (60.9%), information shared by public health authorities (49.1%) and new measures in place (e.g. regarding travel) (42.7%) as main reasons for this change. On the other hand, participants who became less in favor of vaccination did so mainly due to the information shared in the media (51.2%), by public health authorities (41.9%), as well as a change in the sanitary situation (33.7%) (Table 2b).

### Drivers of vaccination intention

Vaccination intention differed by demographic characteristics, with men compared to women (adjusted odds ratio (aOR) 1.37, 95% CI 1.12-1.67) and older adults compared to adults aged 18 to 34 years (aOR 1.86, 95% CI 1.36-2.55, and aOR 5.55, 95% CI 3.79-8.19, for 50-64 and 65 years and older, respectively) more likely to accept COVID-19 vaccination (Table 3).

**Table 3.** Reasons for change in vaccination intention in the past three months.

| Reasons for change in intention | More in favor of vaccination<br>(N=805)<br>n (%) | Less in favor of vaccination<br>(N=86)<br>n (%) |
| --- | --- | --- |
| Change in the sanitary situation | 490 (60.9) | 29 (33.7) |
| Information shared by public health<br>authorities | 395 (49.1) | 36 (41.9) |
| New measures in place (e.g. regarding<br>travel) | 344 (42.7) | 18 (20.9) |
| Scientific developments | 313 (38.9) | 13 (15.1) |
| Information in the media | 190 (23.6) | 44 (51.2) |
| Advice from relatives or friends | 186 (23.1) | 12 (14) |
| Arrival of a new vaccine | 112 (13.9) | 12 (14) |
| Information in social media | 17 (2.1) | 11 (12.8) |

**Table 4.**
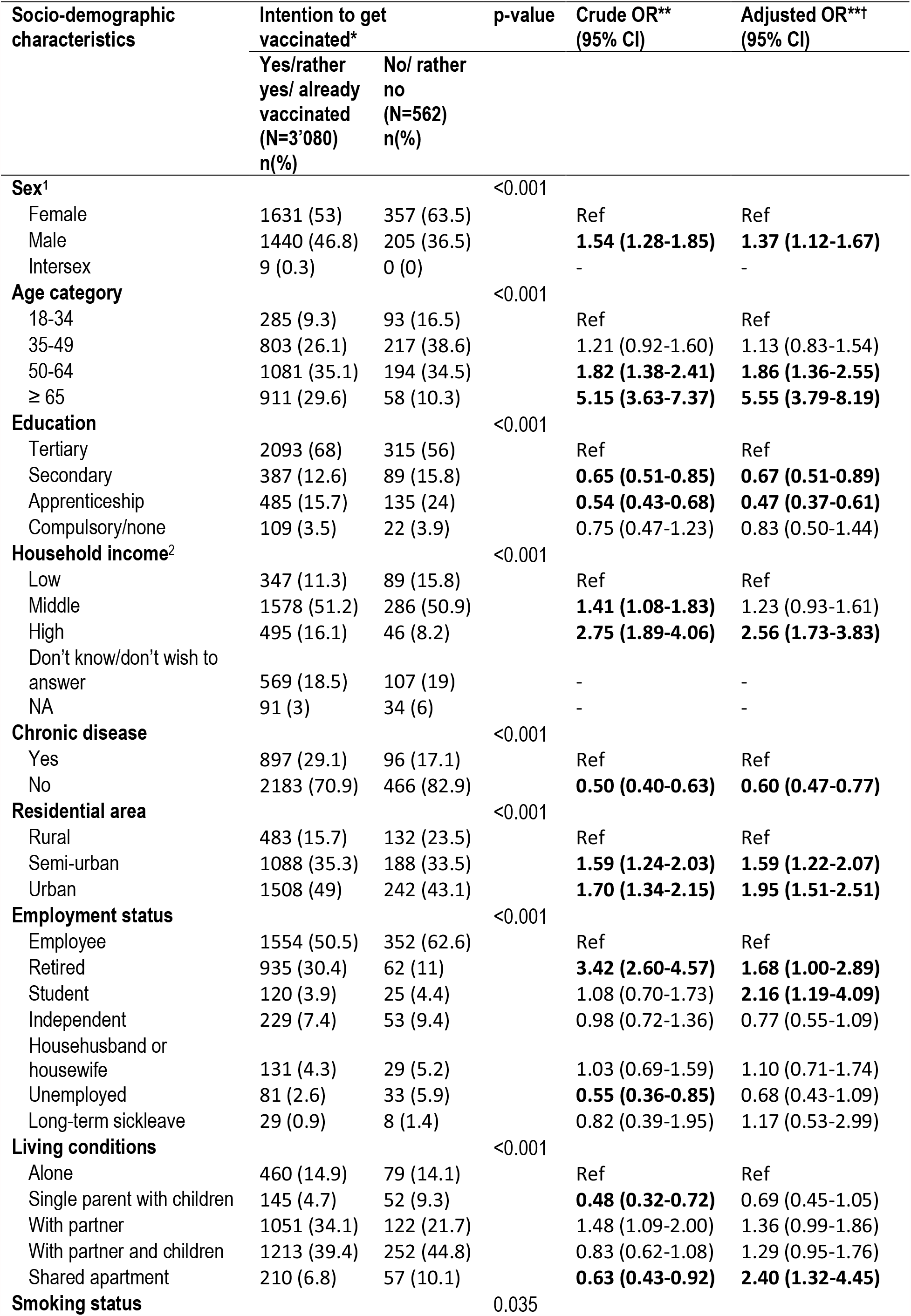
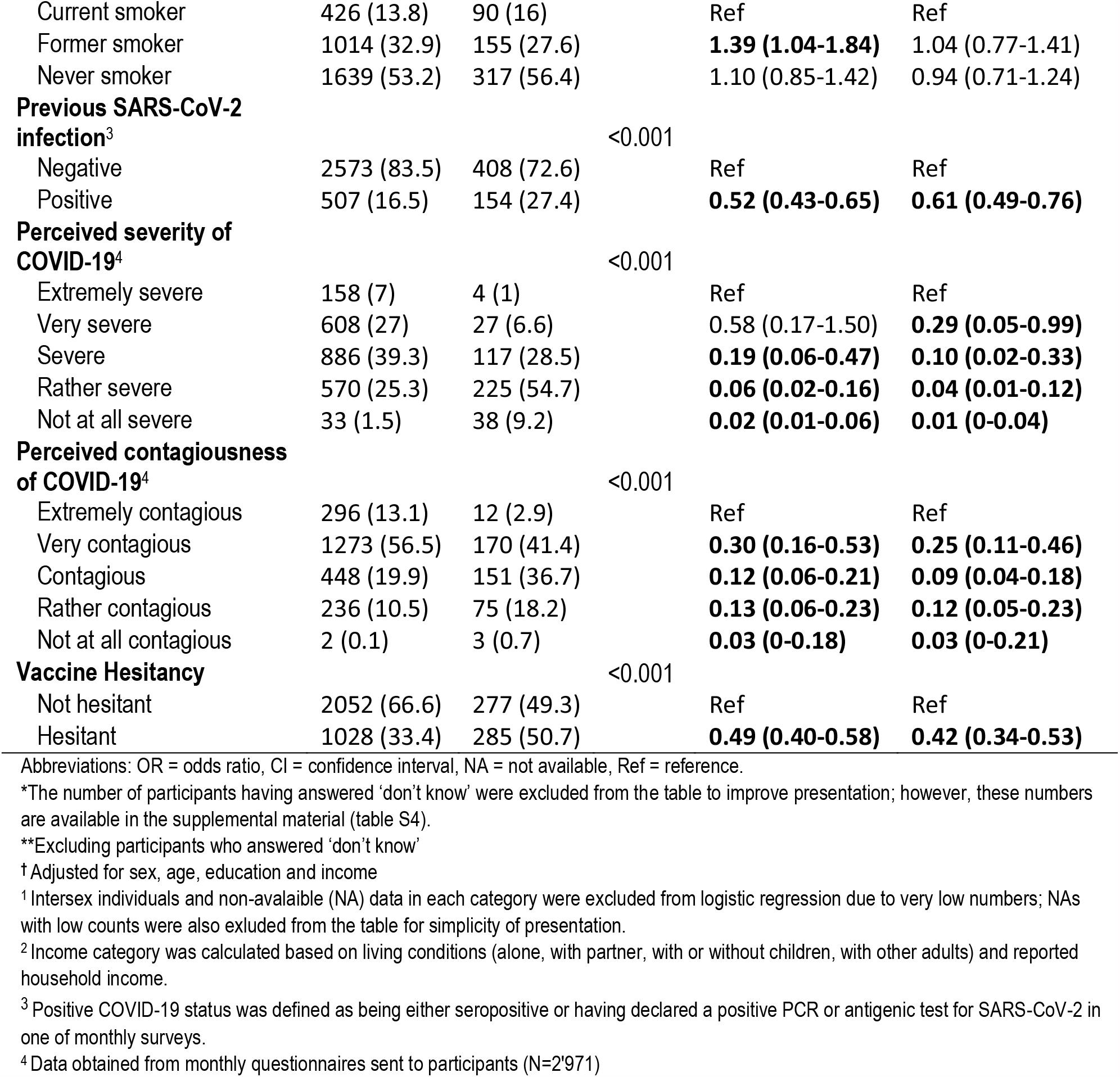
Association of socio-demographic and health-related factors with vaccination intention

Regarding socio-economic characteristics, people who had done apprenticeships (aOR 0.47, 95% CI 0.37-0.61) or had a secondary education (aOR 0.67, 95% CI 0.51-0.89) were less likely to intend to get vaccinated than people having completed tertiary education. The odds did not differ significantly for individuals with compulsory or no formal education. Further, the odds of vaccination willingness were higher in individuals with higher income compared to low income (OR 2.56, 95% CI 1.73-3.83), in retirees (aOR 1.68, 95% CI 1-2.89) and students (aOR 2.16, 95% CI 1.19-4.09) compared to individuals in the workforce, as well as in participants residing in semi-urban (aOR 1.59, 95% CI 1.22-2.07) and urban (aOR 1.95, 95% CI 1.51-2.51) areas compared to rural areas.

Vaccination intention also differed by clinical characteristics, with people without any chronic disease less likely to intend to get vaccinated than those having reported at least one chronic disease (aOR 0.6, 95% CI 0.47-0.77), as well as individuals who had been infected by SARS-CoV-2 compared to those who had never had COVID-19 (aOR 0.61, 95% CI 0.49-0.76).

People who had a lower perception of severity and contagiousness of COVID-19 were less likely to accept vaccination than those with a higher risk perception, with an increasing trend of vaccination intention from those grading COVID-19 as ‘not at all severe’ (aOR 0.01, 95% CI 0-0.04) to ‘very severe’ (aOR 0.29, 95% CI 0.05-0.99) compared to ‘extremely severe’, and from ‘not at all contagious’ (aOR 0.03, 95% CI 0-0.21) to ‘very contagious’ (aOR 0.25, 95% CI 0.11-0.46) compared to ‘extremely contagious’. Finally, vaccine hesitancy was negatively associated with vaccination intention (aOR 0.44, 95% CI 0.36-0.53), although 33.4% of those intending to get vaccinated against COVID-19 were categorized as ‘vaccine hesitant’, while almost half of those not intending to get vaccinated were not generally vaccine hesitant.

## Discussion

This study carried out in the canton of Geneva showed an overall COVID-19 vaccination acceptance of 75.7%, including those already vaccinated, and those who intended or rather intended to get vaccinated once eligible. This rate of vaccine acceptance was consistent with previous studies carried out in other developed countries, such as the United states (67%) ^28^, Japan (62.1%) ^29^, Ireland (65%) and the United Kingdom (between 69% to 86% across studies) ^30,31^. However, the range of vaccine acceptance has been seen to vary widely between countries, from 29.4% reported in a study in Jordan, Kuwait and Saudi Arabia, to 86% in in the United Kingdom.^31^

The most frequently provided reasons for intending to get vaccinated were to protect oneself, to protect the community and to return to a normal life. These reasons were similar to those obtained in other countries.^31^ Interestingly, 80% of those willing to get vaccinated were of the opinion that vaccinated people should continue to follow preventive measures against viral spread. This may reflect a generally higher commitment of these participants to respecting public authority recommendations, which imposed the same preventive measures for vaccinated and non-vaccinated individuals alike at the time of the survey.

In our study, vaccination intentions were different depending on socio-demographic factors. Our results showed that men were more willing to get vaccinated than women. This is in line with previous studies on vaccination acceptance.^31^ Indeed, women have been reported to adopt more negative opinions about vaccination, while men have been reported to perceive a higher risk of the disease and to be less easily influenced by rumors surrounding COVID-19.^31^ Only one study conducted in the United States reported a lower acceptance of COVID-19 vaccination in men compared to women. ^32^

Consistent with previous findings, this study found that older individuals were more willing to get vaccinated against COVID-19 than younger individuals.^31^ This could be attributed to the fact that older individuals are at increased risk of mortality and of severe forms of the disease, and had access to vaccination at the time of the survey. However, this finding could evolve rapidly over time as, since the time of this survey, COVID-19 vaccination has now been made available to all individuals aged over 12 years in Switzerland.

In addition, our study showed that high education level, high income status, as well as having a chronic disease were associated with higher vaccination acceptance, which is in line with previously published studies.^31^ While residing in urban or semi-urban areas was associated with vaccination acceptance in our study, other studies conducted in different settings showed conflicting results regarding residential area.^31^ Although targeted communication directed at clinically vulnerable populations during the vaccination campaign seems to have been successful, with increased vaccination acceptance among participants with a chronic disease, our results suggest that tailored communication strategies should also focus on socially vulnerable populations.^33^

Furthermore, participants who had already been infected with SARS-CoV-2 (assessed either by a serologic test or a PCR-test) were less willing to get vaccinated, even though vaccination was also recommended to previously infected individuals in Switzerland at the time of the survey. To our knowledge, this is the first survey to assess vaccination acceptance in association with serological status. Being aware of these associations may provide guidance for stakeholders and health professionals to target hesitant people and potentially adapt or better explain vaccination strategies.

While 14% of participants in the current study expressed unwillingness to get vaccinated, 10% of participants remained undecided regarding their vaccination intention. Making up almost one fourth of all participants, this combined group represents a threat for the success of vaccination campaigns against COVID-19 and the achievement of a high immunization coverage. The main reasons for participants’ refusal of vaccination were concerns about safety and efficacy, and a large proportion of those not intending to get vaccinated reported preferring to wait to have more data about potential side-effects, including in the long term. Previous studies carried out worldwide have also identified doubts about the vaccine’s efficacy and safety among the main reasons for vaccine hesitancy.^31,34^, High media coverage of the vaccination campaign and increased use of social networks may have further fueled controversies such as the potential risk of thromboembolic events following vaccination, with a heavy impact on vaccination acceptance.^35^

Our results show that, at the time of the survey, less than half of participants with children were willing to have their children vaccinated against COVID-19 if it were recommended by public health authorities. This is generally in line with results from other countries^36–39^, which have shown parental acceptance varying between ^36.3%^ in Turkey^38^ to 60.4% in Canada^36^. Consistent with other studies, child vaccination intention varied according to children’s age^35^, with acceptance increasing with the child’s age from 6 years old in our study, and to educational level^36–38^, with a higher acceptance rate among parents with a tertiary education and those with a compulsory education only. Importantly, COVID-19 vaccination was not yet authorized in children under 16 years old at the time of the survey, while it is now recommended for those aged 12 years and older in Switzerland, which may strongly impact parental acceptance. Also, willingness to vaccinate one’s children may increase as more evidence is being made available regarding potential long-term sequelae of COVID-19 in children and adolescents.^40^

In our study, 33.4% of those intending to get vaccinated against COVID-19 or already vaccinated were categorized as generally ‘vaccine hesitant’, while almost half of those not intending to get vaccinated were not generally vaccine hesitant. This result suggests that COVID-19 vaccination does not seem to be perceived in the same way as other recommended vaccines. Although it is already known that urgently released vaccines are received with greater skepticism than established or well-known vaccines^33^, COVID-19 vaccines may trigger even higher distrust due to their unusually rapid development.

### Implications for public health policies

Results from this study provide a clear insight of socio-demographic subgroups that remain hesitant or refuse COVID-19 vaccination. Interestingly, more than half of those not intending to get vaccinated against COVID-19 agreed that the vaccine was an important step to end the pandemic. As suggested by our results, these individuals may be more likely to change their minds if reassured about the security of vaccines. Although there is sufficient clinical evidence about the efficacy, safety and side effects of authorized COVID-19 vaccines, this evidence needs to be better communicated and disseminated among the general population in order to alleviate common concerns.

Further, our results showed that information shared by public health authorities could lead to change in intention both in favor and against vaccination in similar proportions. These results highlight the importance of improving communication at a population level. Fortunately, empirical data showed that building vaccination trust among hesitant individuals is possible with effective communication strategies, which could be based on social marketing campaigns at the population level^34,41^ or on targeted campaigns tailored to specific subgroups.^42^ Accordingly, public health organizations, health care professionals and digital platforms should collaborate to guarantee the availability of accessible and accurate information.

### Strengths and limitations

The main strengths of this study are is its large sample size with all adult ages represented, as well as the availability of data on sociodemographic (age, sex, education level, income) and health-related characteristics (serologic status, chronic diseases, smoking status) which allowed stratification of COVID-19 vaccination acceptance according to these factors. Very little research has been conducted on the drivers of COVID-19 vaccine acceptance now that vaccination is available to the general population. Indeed, most previous studies carried out globally were conducted in periods when COVID-19 vaccines were not available or accessible only to certain groups, such as healthcare workers or key workers.^43,44^ It is also of outright importance to investigate the factors influencing perception of COVID-19 vaccination at the local level, as vaccination hesitancy may be widely influenced by regional and cultural factors.

Several limitations of our study should be acknowledged. Although participation rate in this study was high, generalization of the results presented here requires caution as our sample is not completely representative of the general population of the canton of Geneva. While we mitigated this by collecting and adjusting our results for important socio-demographic characteristics, a selection bias remains. Participation required French literacy, internet access and digital literacy, potentially excluding part of the general population. Respondents to the survey were older than the general population in Geneva. As older individuals are more vulnerable to COVID-19, this may have resulted in an overestimate of the overall acceptance of COVID-19 vaccination.

Furthermore, the cross-sectional survey design represents a snapshot in time, rather than the evolving landscape of the public’s attitudes about COVID-19 vaccination. Vaccine hesitancy, perceptions, and concerns may change over time. Our results should be interpreted considering this specific time period when vaccination was only accessible to people aged above 65 years or to people with chronic diseases at risk of severe forms of COVID-19. Our survey will be repeated over time to provide updated information and adjust public health messages as appropriate. Furthermore, other factors potentially impacting vaccination hesitancy were not investigated, such as origin, religious or political views, which are likely to influence individual perceptions and behavior. Another aspect that could be worth exploring is the ‘imitation effect’ or the influence of one’s social network on vaccination perception.

Finally, we need to keep in mind that acceptance or intent does not automatically translate into actual behavior. Despite a high vaccination rate to date, with more than half of the population of the canton of Geneva vaccinated with at least one dose45, there is a risk that the vaccination rate could reach a plateau, especially in the context of summer holidays and the slowdown of the pandemic in past months. Once vaccination against COVID-19 of all willing individuals will have been achieved, increased efforts will have to be put in place to reach the more reluctant part of the population and those with fewer access to information about the vaccination campaign.

## Conclusion

Our study found that 75.7% of our sample from the canton of Geneva would accept COVID-19 vaccination or was already vaccinated at the time of the survey. However, socio-demographic variations in rates of acceptance were evidenced that need to be carefully addressed. Policy makers and stakeholders should provide reassuring messages about side effects and effectiveness of the vaccination. This cross-sectional survey will be repeated approximately every six months in order to follow the level of COVID-19 vaccination acceptance over time, which may be influenced by new incentives such as the establishment of a COVID-19 vaccine certificate or new policy for traveling, as well as the pandemic progression and new outbreaks. These data may help inform policy makers to develop effective and targeting communication strategies.

## Data Availability

The data used within this study is made available upon reasonable request to the last author.

## Authors’ contributions

SS, IG and HB designed the study. HB, AW, MN and SS designed the questionnaire for the survey. MZ, FP, HB and AW were involved in participant recruitment and implementation of the survey. NP conducted statistical analyses of the data. AW and HB drafted the manuscript. All authors participated to analysis interpretation and approved the final manuscript.

## Acknowledgements

We thank all the participants, without whom this study would not have been possible.

