## supplementary materials for "COVID-19 Vaccination acceptance in the canton of Geneva: A Cross-Sectional Population-Based Study"

#### Part 1: Study questionnaire

The following questions were included in the analysis (in French in the original questionnaire):

(1) Have you already been vaccinated with at least one dose of the vaccine against COVID-19? (Yes / No / I have an appointment for the first dose)
(2) Once you will be eligible for the COVID-19 vaccination, do you intend to get vaccinated? (Yes / Rather yes / Rather no / No).
(3) Then, participants had to clarify the reasons why they would get vaccinated or not (with a list of possible answers, including ‘other’ entered as free text).
(4) For those (rather) not intending to get vaccinated: what would make you change your mind in favor of vaccination? (list of possible answers, including ‘other’ entered as free text)
(5) Do you think that the vaccine is an important step to end the pandemic?
(6) Do you think that vaccinated people should continue to adopt preventive measures (such as wearing a mask)?
(7) In the past three months, have you changed your mind about COVID-19 vaccination? If so, participants were asked to explain what had changed their mind (list of possible answers, including ‘other’ entered as free text).
(8) If vaccination against COVID-19 were recommended to children, would you be willing to vaccinate your children? (Yes / Rather yes / I don’t know / Rather no / No / I do not have children or my children are above 18)

The questionnaire also included three questions from a French study on vaccination hesitancy[1] adapted from the World Health Organization’s Strategic Advisory Group of Experts (SAGE) definition of vaccine hesitancy[2]:

(1) Have you ever refused, (for your child or yourself), a vaccine recommended by your physician, because you considered this vaccination as dangerous or useless?
(2) Have you ever delayed a vaccine recommended by your physician, for your child or yourself because you hesitated over it?
(3) Have you ever had a vaccine, (for your child or yourself), despite having doubts about its effectiveness?

With possible answers being ‘Yes’, ‘No’, ‘I Don’t know’.

### Part 2: Supplementary Figures and Tables

**Figure S1.** Flow chart of the Specchio-COVID19 Vaccination survey

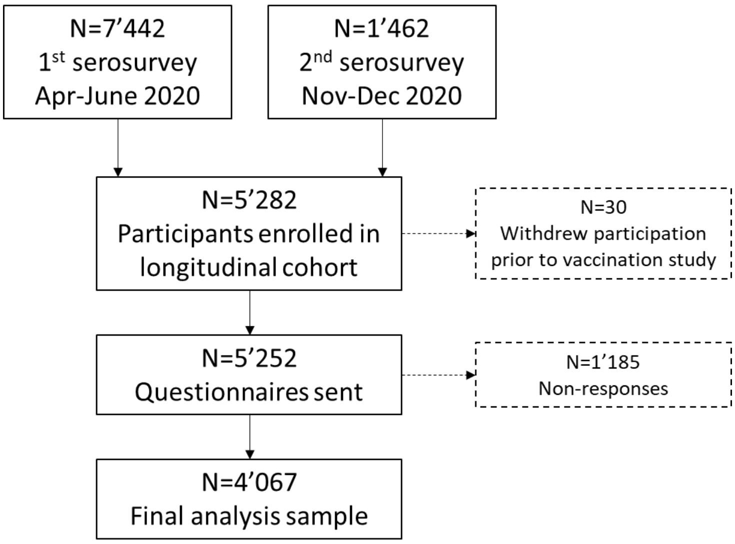

**Table S1** Socio-demographic and health-related characteristics of respondents (n=4'067) vs. non-respondents (n=1'185) to the vaccination questionnaire

| Socio-demographic characteristics | Respondents n (%) | Non-respondents n (%) | p-value |
|---|---|---|---|
| **Sex** | | | 0.002 |
| Female | 2276 (56) | 626 (52.8) | |
| Male | 1780 (43.8) | 548 (46.2) | |
| Intersex | 11 (0.3) | 11 (0.9%) | |
| **Mean age +/- SD** | 53.3 +/- 14.4 | 43.8 +/- 14.4 | |
| **Age category** | | | < 0.001 |
| 18-34 | 423 (10.4) | 299 (25.2) | |
| 35-49 | 1184 (29.1) | 511 (43.1) | |
| 50-64 | 1439 (35.4) | 261 (22) | |
| ≥ 65 | 1021 (25.1) | 114 (9.6) | |
| **Education** | | | < 0.001 |
| Compulsory/none | 158 (3.9) | 81 (6.8) | |
| Apprenticeship | 722 (17.8) | 160 (13.5) | |
| Secondary | 546 (13.4) | 193 (16.3) | |
| Tertiary | 2631 (64.7) | 749 (63.2) | |
| **Household income**[1] | | | < 0.001 |
| Low | 523 (12.9) | 193 (16.3) | |
| Middle | 2055 (50.5) | 498 (42) | |
| High | 582 (14.3) | 164 (13.9) | |
| Don't know/don't wish to answer | 773 (19) | 258 (21.8) | |
| **Chronic disease** | | | < 0.001 |
| Yes | 1094 (26.9) | 227 (19.2) | |
| No | 2973 (73.1) | 956 (80.7) | |
| **Employment status** | | | < 0.001 |
| Employee | 2174 (53.5) | 726 (61.3) | |
| Retired | 1052 (25.9) | 116 (9.8) | |
| Student | 162 (4) | 142 (12) | |
| Independent | 320 (7.9) | 79 (6.7) | |
| Househusband or housewife | 183 (4.5) | 62 (5.2) | |
| Unemployed | 132 (3.2) | 46 (3.9) | |
| Long-term sick leave | 43 (1.1) | 14 (1.2) | |
| **Living conditions** | | | < 0.001 |
| Alone | 589 (14.5) | 110 (9.3) | |
| Single parent with children | 238 (5.9) | 68 (5.7) | |
| With partner | 1264 (31.1) | 195 (16.5) | |
| With partner and children | 1675 (41.2) | 623 (52.6) | |
| Shared apartment | 300 (7.4) | 189 (15.9) | |
| **Smoking status** | | | < 0.001 |
| Current smoker | 606 (14.9) | 227 (19.2) | |
| Former smoker | 1296 (31.9) | 282 (23.8) | |
| Never smoker | 2164 (53.2) | 676 (57) | |
| **Previous SARS-CoV-2 infection**[3] | | | 0.005 |
| Positive | 772 (19) | 269 (22.7) | |
| Negative | 3295 (81) | 916 (77.3) | |

SD: standard deviation.
[1] Income category was calculated based on living conditions (alone, with partner, with or without children, with other adults) and reported household income.
[2] Previous SARS-CoV-2 infection was defined as being either seropositive or having declared a positive PCR or antigenic test for SARS-CoV-2 in one of monthly surveys.

**Table S2.** Comparison of education level and age category between the Canton of Geneva and the vaccination survey respondents.

| | Women | | Men | | Overall | |
|---|---|---|---|---|---|---|
| | Geneva N (%) | Survey respondents N (%) | Geneva N (%) | Survey respondents N (%) | Geneva N (%) | Survey respondents** N (%) |
| **Education level** | | | | | | |
| Tertiary | 77117 (20.1) | 1404 (34.7) | 76217 (19.9) | 1220 (30.2) | 153334 (39.9) | 2624 (64.9) |
| Secondary* | 65639 (17.1) | 766 (18.9) | 59500 (15.5) | 498 (12.3) | 125139 (32.6) | 1264 (31.2) |
| Compulsory | 57509 (15) | 99 (2.4) | 47930 (12.5) | 59 (1.5) | 105439 (27.5) | 158 (3.9) |
| **Age category** | | | | | | |
| 18-34 | 58026 (14) | 266 (6.6) | 57421 (13.9) | 155 (3.8) | 115447 (27.9) | 421 (10.4) |
| 35-49 | 58556 (14.1) | 712 (17.6) | 56718 (13.7) | 470 (11.6) | 115274 (27.8) | 1182 (29.1) |
| 50-64 | 50892 (12.3) | 802 (19.8) | 48949 (11.8) | 632 (15.6) | 99841 (24.1) | 1434 (35.4) |
| ≥ 65 | 48509 (11.7) | 496 (12.2) | 35303 (8.5) | 523 (12.9) | 83812 (20.2) | 1019 (25.1) |

*Secondary education level combines ‘apprenticeship’ and ‘secondary’ education from Table S1.
**Intersex individuals were not included in this table due to lack of data on this category in the statistics of the canton of Geneva.

**Table S3.** Willingness to vaccinate one's children (N=1'339) stratified by parents' vaccination, education level and child's age.

| | Willingness to vaccinate one's children | | | | |
|---|---|---|---|---|---|
| | Yes | Rather yes | Don't know | Rather no | No |
| **Parent's vaccination intention, N (%)** | | | | | |
| Yes | 341 (37) | 245 (26.6) | 233 (25.3) | 73 (7.9) | 29 (3.1) |
| No | 3 (1.3) | 10 (4.3) | 29 (12.4) | 60 (25.6) | 132 (56.4) |
| Don't know | 2 (1.1) | 10 (5.4) | 88 (47.8) | 47 (25.5) | 37 (20.1) |
| **Parent's education level, N(%)** | | | | | |
| Tertiary | 296 (29.6) | 209 (20.9) | 242 (24.2) | 123 (12.3) | 130 (13) |
| Secondary | 17 (12.8) | 19 (14.3) | 46 (34.6) | 21 (15.8) | 30 (22.6) |
| Apprenticeship | 23 (13.9) | 29 (17.6) | 49 (29.7) | 33 (20) | 31 (18.8) |
| Compulsory/none | 10 (25.6) | 8 (20.5) | 11 (28.2) | 3 (7.7) | 7 (18) |
| **Child's age* (years), N(%)** | | | | | |
| 0-5 | 102 (25.8) | 78 (19.7) | 97 (24.5) | 56 (14.1) | 63 (15.9) |
| 6-10 | 78 (20.7) | 67 (17.8) | 120 (31.9) | 51 (13.6) | 60 (16) |
| 11-15 | 110 (27.1) | 86 (21.2) | 98 (24.1) | 52 (12.8) | 60 (14.8) |
| 16-18 | 56 (34.8) | 34 (21.1) | 35 (21.7) | 21 (13) | 15 (9.3) |

*When a participant had several children, the age of the youngest child was considered in the categorization.

**Table S4.** Socio-demographic and health-related characteristics, and risk perception stratified by intention of vaccination against COVID-19.

| Characteristics | Intention to get vaccinated | | |
|---|---|---|---|
| | Yes/rather yes/ already vaccinated (N=3'080) n(%) | No/ rather no (N=562) n(%) | Don't know (N=425) n(%) |
| **Sex** | | | |
| Female | 1631 (53) | 357 (63.5) | 288 (67.8) |
| Male | 1440 (46.8) | 205 (36.5) | 135 (31.8) |
| Intersex | 9 (0.3) | 0 (0) | 2 (0.5) |
| **Age category** | | | |
| 18-34 | 285 (9.3) | 93 (16.5) | 45 (10.6) |
| 35-49 | 803 (26.1) | 217 (38.6) | 164 (38.6) |
| 50-64 | 1081 (35.1) | 194 (34.5) | 164 (38.6) |
| ≥ 65 | 911 (29.6) | 58 (10.3) | 52 (12.2) |
| **Education** | | | |
| Tertiary | 2093 (68) | 315 (56) | 223 (52.5) |
| Secondary | 387 (12.6) | 89 (15.8) | 70 (16.5) |
| Apprenticeship | 485 (15.7) | 135 (24) | 102 (24) |
| Compulsory/none | 109 (3.5) | 22 (3.9) | 27 (6.4) |
| **Household income**[1] | | | |
| Low | 347 (11.3) | 89 (15.8) | 87 (20.5) |
| Middle | 1578 (51.2) | 286 (50.9) | 191 (44.9) |
| High | 495 (16.1) | 46 (8.2) | 41 (9.6) |
| Don't know/don't wish to answer | 569 (18.5) | 107 (19) | 97 (22.8) |
| NA | 91 (3) | 34 (6) | 9 (2.1) |
| **Chronic disease** | | | |
| Yes | 897 (29.1) | 96 (17.1) | 101 (23.8) |
| No | 2183 (70.9) | 466 (82.9) | 324 (76.2) |
| **Residential area** | | | |
| Rural | 483 (15.7) | 132 (23.5) | 72 (16.9) |
| Semi-urban | 1088 (35.3) | 188 (33.5) | 161 (37.9) |
| Urban | 1508 (49) | 242 (43.1) | 192 (45.2) |
| **Employment status** | | | |
| Employee | 1554 (50.5) | 352 (62.6) | 268 (63.1) |
| Retired | 935 (30.4) | 62 (11) | 55 (12.9) |
| Student | 120 (3.9) | 25 (4.4) | 17 (4) |
| Independent | 229 (7.4) | 53 (9.4) | 38 (8.9) |
| Househusband or housewife | 131 (4.3) | 29 (5.2) | 23 (5.4) |
| Unemployed | 81 (2.6) | 33 (5.9) | 18 (4.2) |
| Long-term sickleave | 29 (0.9) | 8 (1.4) | 6 (1.4) |
| **Living conditions** | | | |
| Alone | 460 (14.9) | 79 (14.1) | 50 (11.8) |
| Single parent with children | 145 (4.7) | 52 (9.3) | 41 (9.6) |
| With partner | 1051 (34.1) | 122 (21.7) | 91 (21.4) |
| With partner and children | 1213 (39.4) | 252 (44.8) | 210 (49.4) |
| Shared apartment | 210 (6.8) | 57 (10.1) | 33 (7.8) |
| **Smoking status** | | | |
| Current smoker | 426 (13.8) | 90 (16) | 90 (21.2) |
| Former smoker | 1014 (32.9) | 155 (27.6) | 127 (29.9) |
| Never smoker | 1639 (53.2) | 317 (56.4) | 208 (48.9) |

| | | | |
|---|---|---|---|
| **Previous SARS-CoV-2 infection**[2] | | | |
| Negative | 2573 (83.5) | 408 (72.6) | 314 (73.9) |
| Positive | 507 (16.5) | 154 (27.4) | 111 (26.1) |
| **Perceived severity of COVID-19**[3] | | | |
| Extremely severe | 158 (7) | 4 (1) | 8 (2.6) |
| Very severe | 608 (27) | 27 (6.6) | 36 (11.8) |
| Severe | 886 (39.3) | 117 (28.5) | 125 (41) |
| Rather severe | 570 (25.3) | 225 (54.7) | 129 (42.3) |
| Not at all severe | 33 (1.5) | 38 (9.2) | 7 (2.3) |
| **Perceived contagiousness of COVID-19**[3] | | | |
| Extremely contagious | 296 (13.1) | 12 (2.9) | 18 (5.9) |
| Very contagious | 1273 (56.5) | 170 (41.4) | 149 (48.9) |
| Contagious | 448 (19.9) | 151 (36.7) | 96 (31.5) |
| Rather contagious | 236 (10.5) | 75 (18.2) | 42 (13.8) |
| Not at all contagious | 2 (0.1) | 3 (0.7) | 0 (0) |

NA (not available) results with low counts were excluded from the table for simplicity of presentation.

[1] Income category was calculated based on living conditions (alone, with partner, with or without children, with other adults) and reported household income.

[2] Positive COVID-19 status was defined as being either seropositive or having declared a positive PCR or antigenic test for SARS-CoV-2 in one of monthly surveys.

[3] Data obtained from monthly questionnaires sent to participants (N=2'971)
